# Improving Quality of CAR-T Cell Therapy Starting Material with Automated Microfluidic Cell Sorting

**DOI:** 10.64898/2025.12.16.25342401

**Authors:** Alison M. Skelley, Yasna Behmardi, Luke Petersen, Mabel Shehada, Laurissa Ouaguia, Khushroo Gandhi, Roberto Campos-González, Tony Ward

## Abstract

Autologous CAR-T cell therapy has demonstrated remarkable clinical efficacy in hematologic malignancies, yet its broader application remains limited by complex, labor-intensive manufacturing and inconsistent product quality. We describe a novel microfluidic cell separation platform based on Deterministic Lateral Displacement (DLD), integrated into a fully automated, closed-system instrument (Curate System), capable of processing full leukopacks in under one hour. Compared to Ficoll®-based density gradient centrifugation, DLD processing yielded significantly higher leukocyte recovery (88% vs. 58%), superior platelet and red blood cell depletion, and reduced CD69⁺ T-cell activation. Flow cytometric analysis revealed improved phenotypic preservation across key T-cell subsets, including naïve and central memory populations. Cytokine profiling demonstrated enhanced washing efficiency, with markedly lower levels of biologic response modifiers such as RANTES and TGF-β1. DLD-purified T cells exhibited enhanced expansion kinetics and greater yield, supporting improved manufacturing outcomes. These findings position DLD-based processing as a clinically relevant, scalable alternative to conventional methods, with potential to improve consistency, potency, and accessibility of CAR-T therapies.

## INTRODUCTION

Autologous cell therapy has seen accelerated progress towards eliminating blood cancers using patient derived engineered T cells. Unfortunately, the reality is that current therapies remain far from accessible for most patients, with costs that can reach upwards of $500,000.^1–3^ Despite therapies being available since 2017, and with 9 therapies currently on the market, fewer than 40,000 patients have been treated.^4^ One major hurdle is the very manual and lengthy manufacturing process. Patient leukapheresis collections frequently contain the minimum number T cells to create multiple therapeutic doses, but due to the inefficiencies of downstream processes multiple days of *ex-vivo* expansion are required to achieve the desired number of cells.^5^ Several steps in the process require manual manipulation of the sample, and manufacturing facilities commonly do not have capacity or staff to manufacture more than a few therapies at a time.^6^ As a result, strict evaluation criteria and long wait times – upwards of 6 months – are a common reality.^6^ Biological variability in starting materials and complex manufacturing processes result in inconsistent therapeutic outcomes; furthermore up to 4-7% manufacturing processes fail and patients never receive their therapy.^7^ From a clinical perspective, cell therapy is often a therapy of last resort, and yet a significant portion of patients may require additional apheresis collections, bridging therapy, or because of a failed manufacturing process may not get their therapy at all.^8,9^

Widespread adoption of CAR-T therapies is further complicated by mounting evidence suggesting cell quality has a significant impact on the treatment potency and efficacy. Recent clinical trials have shown numerous contributing factors.^7^ Patients who had apheresis collections at the time of diagnosis had better outcomes than those who were waitlisted.^10,11^ Lower levels of functional exhaustion markers in apheresis starting material corresponded with a higher likelihood of therapeutic success.^12^ Consistent expansion of cells vs. a lag in proliferation resulted in a better patient response.^13^ Conversely, longer *ex vivo* culture time was associated with differentiated T-cells and depletion of stem cell memory (T_scm_) and naïve phenotypes,^14^ potentially leading to poor efficacy due to T-cell exhaustion and apoptosis.^15^ Higher percentages of central memory T-cells (T_CM_)^16,17^ and lower levels of regulator T-cells (T_REG_)^18^ resulted in greater *in vivo* persistence and greater anti-tumor activity. Patients who had greater *in vivo* persistence of CAR-T cells had improved long-term clinical outcomes.^16,19^ While alleviation of manufacturing hurdles and cost reduction is a primary focus, careful attention must be given to optimizing cell quality both into and out of the manufacturing process in order to achieve the best therapeutic outcome.

Recently, great strides have been made to improve the CAR-T process, including generating CAR-T constructs that have greater anti-tumor effects and *in vivo* persistence,^20^ and developing methods that require fewer days of expansion to reach dose.^14,21,22^ Some exciting studies propose methods with no *ex vivo* expansion required at all.^23,24^ In addition, numerous systems launched in the last few years have focused on “all-in-one” processing, with automated workflows that significantly reduce manual labor and promise greater efficiency in the downstream processes (Miltenyi CliniMACS Prodigy®, Lonza Cocoon®, Cellares Cell Shuttle™, and Ori Biotech IRO^®^ to name a few). These advancements potentially enable shorter vein-to-vein time, minimize the need for bridging therapy and increased life expectancy.^25^ Furthermore, higher capacity at manufacturing facilities can help bring down costs by relying on automation instead of costly manual labor, while increasing the number of patients that can be treated. With shorter processes, however, comes even greater risk that contaminating cells and other materials in the starting material still remain after expansion, and are dosed back to the patient.^26^ This can not only limit the efficacy of the therapy, but can cause adverse responses such as Cytokine Release Syndrome (CRS) and immune effector cell-associated neurotoxicity syndrome (ICANS).^26–28^ The cell therapy field requires a robust, consistent, automated and efficient cell preparation method that can deliver the high cell counts, high purity and high quality needed for input into these new rapid processes.

In typical CAR-T cell manufacturing an initial debulking step is performed post-apheresis collection prior to T-cell selection and activation. Commonly used systems include density gradient centrifugation (Sepax™ C-Pro, Cytiva), counterflow centrifugation (Gibco™ CTS™ Rotea™, Thermo Fisher Scientific), and spinning membrane instruments (Lovo^®^, Fresenius Kabi). Although these instruments recover WBC they do so with variable cell losses, and with variable removal of erythrocytes and/or platelets. Contamination with platelets can alter the T cells and suppress proliferation during culture.^29^ It is also widely known that centrifugation induces stress and non-desirable cell to cell interactions.^30–32^ Additional treatment with ammonium chloride to lyse remaining erythrocytes can also have negative impact on cell quality^32–34^.

Previously, we described a microfluidic chip that used the principles of Deterministic Lateral Displacement (DLD) to process blood and blood products for the production of CAR-T cells.^35,36^ This proof-of concept scale device demonstrated superior purification and phenotypic advantages in the DLD product compared to density gradient centrifugation. In this study, we focus on the next generation of cell separation devices, optimized and scaled up to process full leukopacks (volumes of up to 300mL) within an hour. We also describe the integration of these massively parallel microfluidic arrays into a novel fully closed single-use disposable cassette and its use on an easy-to-use automated prototype cell processing system. We characterize the performance of this system through head-to-head runs with density gradient centrifugation (one of the most widely used methods for debulking post-apheresis) and show the functional benefits of cells isolated using the DCS approach.

## MATERIALS AND METHODS

### Microfluidic DCS device and cassette fabrication

DCS microarrays were constructed in ZEONEX^®^ Cyclo Olefin Polymer (COP, Zeon Corporation) via compression molding (Edge Precision Manufacturing, a Zeon Corporation subsidiary). Molded arrays were lidded with COP film. Manifold layers were prepared by heat staking hydrophobic vents at the terminus of each manifold channel. These vents facilitated air leaving during the priming operation. Paired lidded elements were laser welded on each side of the central manifold layer. Single barbed tubing connections were made for each of the 4 manifold channels (sample, running buffer, product and waste). Two manifold units (net 4x microfluidic layers) were connected together via barbed y-connectors and then connected to the cassette via a single set of inputs and outputs. The cassettes were created by laser welding a thin film to front and back side of an injection molded cassette body (FastRadius). Compression-molded gaskets were placed into the cassette and held in place using a laser-welded gasket frame. Tubing to connect to the various reagent and sample/product/waste bags was secured to the cassette via UV glue. Tubes terminated either in a bag spike or with a sealed end. The sample tube featured an in-line capsule filter with a 20 µm nylon membrane (Clear Solutions). Peristaltic tubing was connected to the barbed pump tube connectors on board the cassette. The fully closed cassette was sterilized using Ethylene Oxide prior to use. The cassette was designed for simple and fast assembling and loading on to the instrument. From connecting sample and reagents to the cassette to starting the run, users with very little training could complete the process in under 15 min.

#### Curate system

The Curate instrument was designed to accommodate the fully closed cassette, using peristaltic pumps to drive flow within the array (Figure 1 A, B). To minimize pulsatile flow, critical for establishing a stable separation profile in the array, each pump featured dual opposed rotors. Briefly, a single fluid line was split into two paths, travelled over two rollers 180 degrees out of phase, and then was recombined, delivering smoother flow. The system contained 4 independent pumps for sample, diluent (allowing in-line dilution), buffer, and waste. Independent control enabled precise management of flow rates, relative ratios and fluid viscosities to establish optimized DLD operations for apheresis processing. No per-run adjustment was used for the data presented in this article.

**Figure 1:**
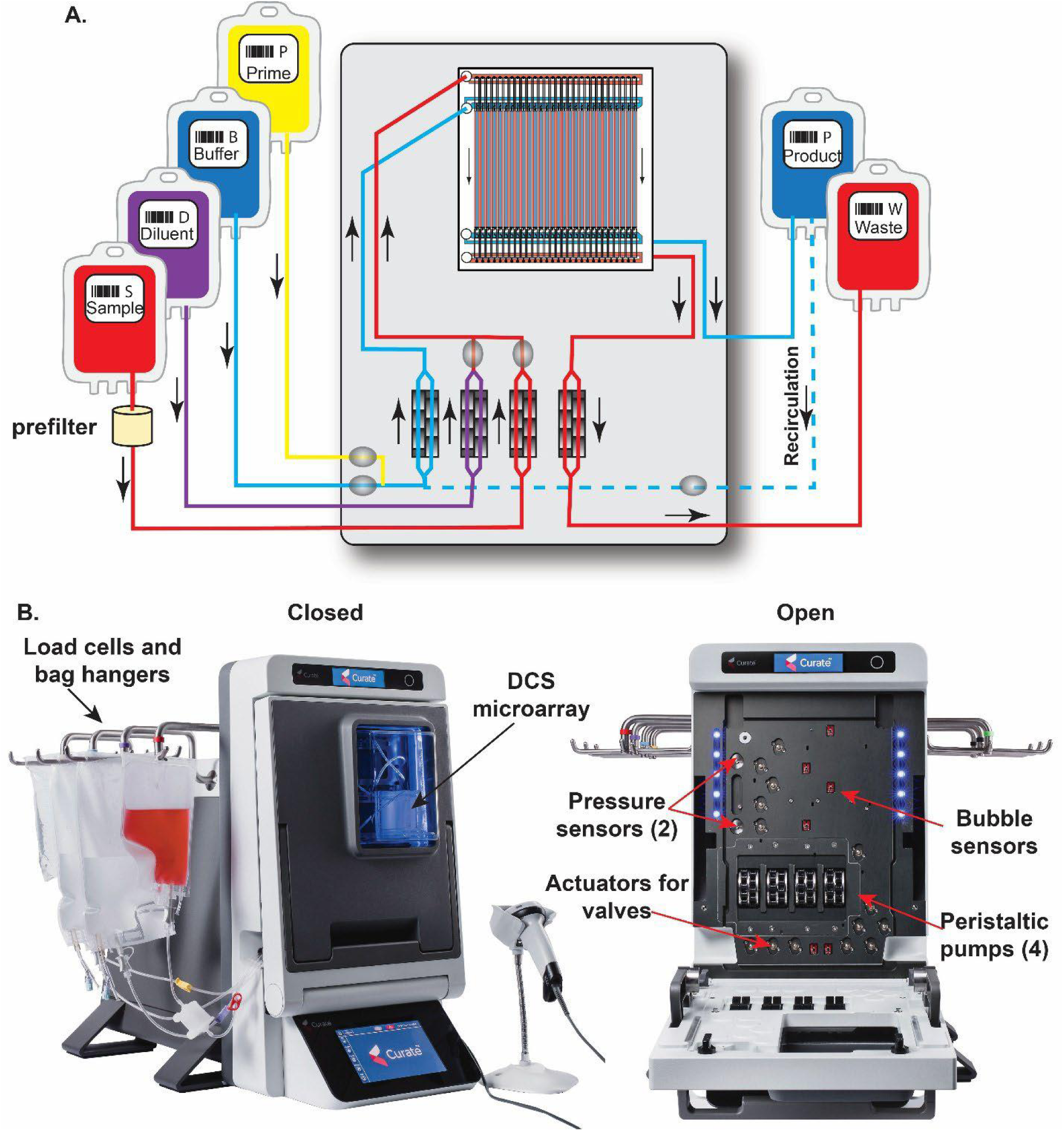
The Curate system for automated microfluidic processing of apheresis samples. A) The fully closed cassette contains a microfluidic array as well as channels, valves and peristaltic tubing to direct fluid into and out of the separation unit. The Curate system (B) operates the cassette via peristaltic pumps and solenoid valves while monitoring separation progress via load cells on the hangers and in-line pressure and bubble sensors.

Fluid flow was controlled by a series of solenoid valves aligned with gaskets on the cassette. Bags for sample, product, and waste were hung on load cell equipped hangars, which verified fluid movements in and out of the DCS array. Pressure and bubble sensors upstream of the microarray were used to monitor and trigger adjustments to maintain fluid ratios as needed.

#### Running buffers

DCD microarrays were primed using 0.6% F-127 in saline. A priming concentrate of 10% F-127 in water was sterile filtered and then bottled; these bottles were then autoclaved prior to storage at 4C. Before the run 15 mL of the concentrate was withdrawn from the bottle and injected into a 250 mL bag of sterile saline (VWR, cat. no. 95037-334) which was then connected to the cassette.

Running buffers commonly consisted of an albumin mixture in phosphate-buffered saline (PBS) or Plasma-Lyte^®^-A Injection, pH 7.4 (PLA, VWR cat. no. 80089-818). Concentrated bovine serum albumin (BSA)or human serum albumin (HSA, Grifols Albumin (Human) 25%, USP, Plasbumin^®^-25, cat. no. 13533-692-71)) was injected into bags of sterile PLA or added to PBS and sterile filtered. In some instances, cell culture media was used, such as TexMACS™ (Miltenyi Biotec) or CTS™ OpTmizer™ (Thermo Fisher Scientific). If reagents were prepared more than a day prior to run, they were sterile filtered (0.2 µm pore size) and then loaded into sterile bags. Sterile collection bags for Product and Waste were connected to the cassette prior to use via tube welding.

#### Sample pre-treatment

Leukopack samples were obtained from apheresis of normal donors (San Diego Blood Bank, Red Cross Los Angeles, Stem Express (now CGT Global), or HemaCare). Upon collection leukopacks were spiked with an additional amount of Anticoagulant Citrate Dextrose Solution, Solution A (ACD-A) for a final citrate concentration of 19mM then transported at 4-8 °C. LKPs were stored overnight at 2-8 °C on a Ohaus rocker with constant movement. Before use the LKPs were brought back to room temperature. A benzonase pre-treatment was performed prior to processing the samples.

Benzonase Nuclease HC (final concentration 50 units/mL, Millipore, cat. no. 71205-3) and MgCl_2_ (final concentration 5 mM) were added to the leukopack followed by 1hr incubation at room temperature while rocking. Before processing, EDTA (final concentration 5mM) was added followed by another 15min incubation at room temperature.

#### Manual density gradient separation

Purification of PBMC by Ficoll-Paque^®^ (Cytiva cat. no. 17544202) was performed using the manufacturer’s instructions^37^. Briefly, 10-20 mL of the leukopack was diluted 1:1 in cell culture media and layered on top of an equal volume of Ficoll®. The sample was centrifuged at 400xg for 35min with no brake. The PBMC layer was carefully removed using a pipet. The PBMC layer was washed via resuspending in 50-fold excess media, followed by pelleting by centrifugation at 400xg for 10 min (wash 1) and then 200xg for 10 min (wash 2). The final pellet (PBMC) was resuspended in media to the original volume. This manual process took ∼1.5 hrs to process only 10-20 mL of the initial sample.

#### FACS cell analysis

Phenotypic analysis and growth of cultured T cells was measured at different days post-transduction by mixing the cells in culture, taking an aliquot and then detaching magnetic beads by pipetting and placing the mixture in a magnet for 5min.^38^. Cells were counted with a CellDyn hematology analyzer (Abbott). Staining for flow cytometry was done using 0.5-1.0×10^6^ cells resuspended in 100µl of 0.5% BSA/PBS. Cells were stained with the corresponding panel(s) of antibodies for 30min in the dark, followed by a wash (for the 9-color panels) and then fixed with 1.0% p-formaldehyde (if necessary) before reading in the Novocyte flow cytometer (Agilent Technologies). Data analysis was done with NovoExpress software (Agilent Technologies). For phenotypic analysis we used concentration optimized panels as follows: Apoptosis - Annexin V-FITC, 7-AAD, CD45-BV421, CD3-APC with cells in 0.5% BSA/PBS + 2.0mM CaCl_2_; For %T cell determination - CD3-BV-421, CD4-PerCP, CD45-FITC and CD8-PE; CD45-FITC, CD14-PE, CD3-BV421, CD19-BV-605 with 0.25 ug/tube 7-AAD added for viability; For T cell subtype and activation: CD11b-BV670, CD95-FITC, CD69-BV510, CD3-BV421, CD4-BV650, CD45RA-BV605, CD25-APC, CDD62L-PE/Cy7, CCR7-PE, CD8-APC/Cy7 and 3.0µM DRAQ7; for enumeration of RBC, PLT and WBC in the input, product and waste - CD3-BV-421, CD41-FITC, CD235a-PE and DRAQ5; for T cell senescence status: CD45-RO-BV570, TIM3-BV510, PD1-BV421, CD4-BV650, CD3-APC, KLRG1-PE/Cy7, CCR7-PE, CD8-APC/Cy7 and DRAQ7. Both the T cell subtype and senescence panels were washed by centrifugation prior to addition of DRAQ7.

#### Cytokine Analysis

Cytokines present in the source material and in products of each separation method were quantified. Ficoll® processed samples were collected following the second wash and resuspension. DCS materials were collected immediately following run completion. Aliquots were removed and normalized to ∼10^7^ WBC cells. The aliquots were centrifuged and then the supernatant removed for analysis. Supernatants were frozen into cryo-vials as soon as practical following each separation and supernatant isolation process. Cryopreserved samples were sent to Eve Technologies for a 52-plex Discovery Panel Luminex analysis targeted to a broad range of cytokines, chemokines and cell damage related biomarkers known to be associated with the inflammation cascade. (Analytes are shown in Supplemental Table 1).

#### T-cell stimulation and expansion

Purified cells either from Ficoll® or DCS were counted and the T cell number was determined before activation and culture. Cells were normalized to a concentration of at 10×10^6^/ml in TexMACs media and then were incubated with pre-washed CD3/CD28 Dynabeads (Thermo Fisher Scientific, cat. no. 11161D) at a ratio of 3:1 beads per T cell) for 1h at 37 °C.^38^ After the incubation the beads/cells slurry was placed in a magnet for 5 minutes and then the supernatant was discarded. The slurry was resuspended to a T-cell concentration of 1×10^6^/ml, or desired concentration, in full media containing TexMACs, 10% Fetal Bovine Serum (FBS, Phoenix Scientific, PS-100) or 5% HSA, plus 5ng/ml of each IL-7 and IL-15 (Biolegend cat no. 777704 and 570308), and 1% Penicillin/Streptomycin (Thermo Fisher Scientific cat. no. 15140122). The cells were plated in 6-well GRex plates, or 1L G-Rex containers (Wilson Wolf Manufacturing cat. no. 80240M and RU81100 respectively) and placed at 37 °C, 5% CO_2_ in a humidified incubator.

## RESULTS

### Microarray design

The microfluidic cell sorting system utilizes Deterministic Lateral Displacement (DLD, also called Deterministic Cell Sorting or DCS) in order to separate cells by size. The DCS principle relies on precisely constructed arrays of microposts that are tilted with respect to the major flow direction. The microarray has a critical diameter (Dc) determined by the basic array parameters of array tilt and gap sizes.^39^ As shown in Figure 1A, cells with diameters greater than the critical diameter are deflected and travel along the direction of the array while cells with diameters less than the Dc, along with diffusible species, are unimpeded by the array and flow straight through to the waste. For the device utilized here a Dc of ∼4.2 µm was selected to deflect all white blood cells to the product, while platelets and red blood cells traveled straight through to waste. Red blood cells (RBCs) traveled through the array on their short axis and thus behaved like ∼ 2 µm objects, passing through the array to the waste collection bag.

The design of the micropost array was optimized for throughput, anticipating the high concentrations of cells present in apheresis samples. To improve throughput per lane we modified the micropost array design to create an asymmetric DCS array.^40^ Widening the gaps between microposts (perpendicular to the flow direction) reduced the fluidic resistance within the array and had the benefit of making the array more resistant to biofouling. We also minimized shear forces on the cells by using microposts with sharp corners^41^ instead of the round microposts used previously.^42^ To maintain the Dc of ∼4 µm while avoiding unnecessarily small gaps – a challenge for high-volume manufacturing of microfluidic devices - we used microposts with a hexagonal cross-section (20 x 40 µm). This created an asymmetric gap profile but did so by elongation as opposed to constriction (Figure 2B). The minimum gap in the array was 17 µm, well within the manufacturability limits for compression molding.

**Figure 2.**
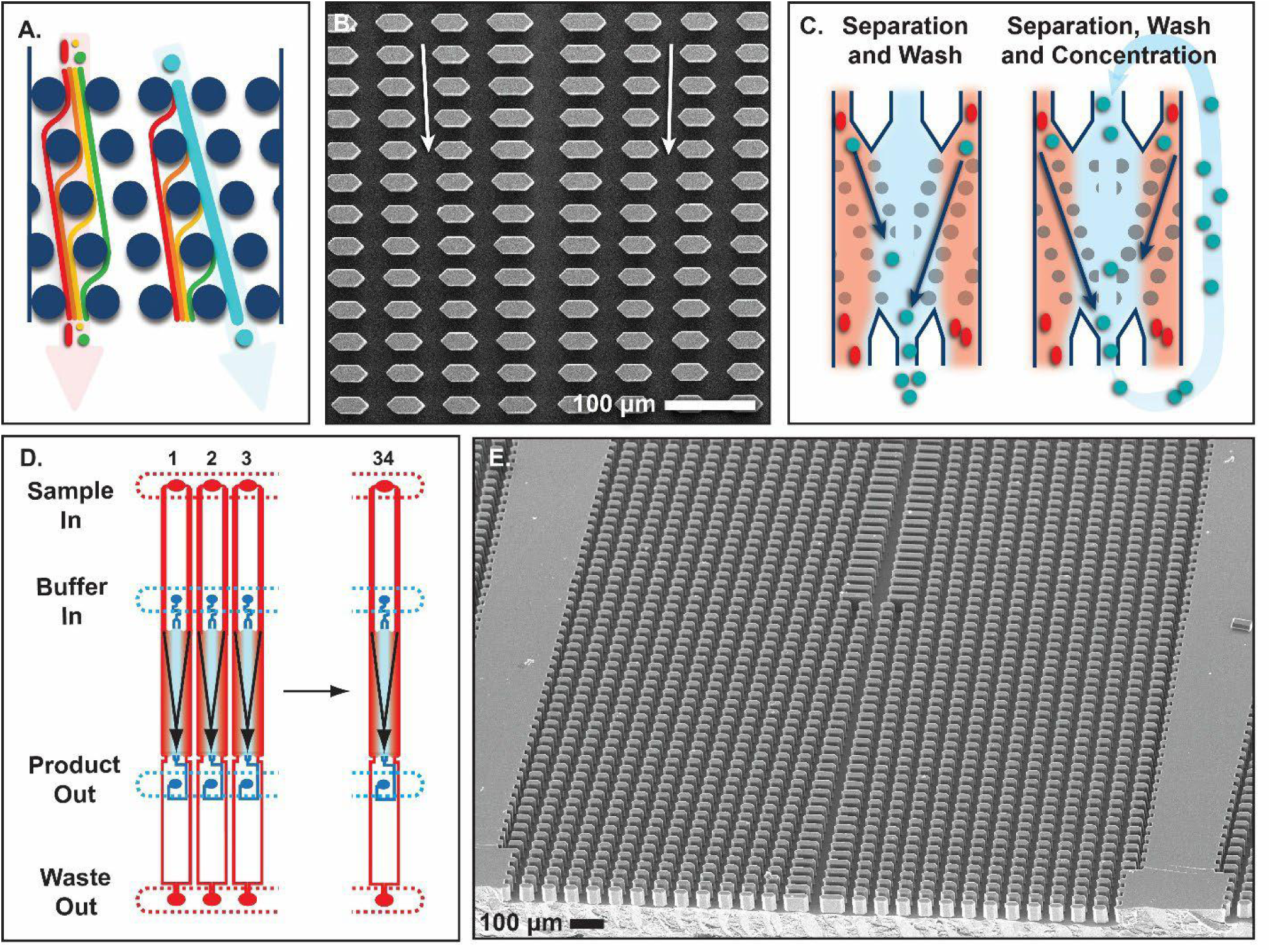
A) Principle of DCS separation. When cells flow through an array that is tilted with respect to the major flow direction they are fractionated by size. Large objects are deflected and follow the array direction while small objects travel straight through. B) SEM image of hexagonal array configuration for lower fluidic resistance and higher throughput. Arrows indicate direction of flow. C) Modes of operation within the microfluidic array. D) Increasing throughput by using multiple lanes in one microfluidic layer. E) SEM image of one array with adjacent arrays (lanes) visible.

In order to both separate and wash the cells into a clean collection buffer, each array contained co-flowing streams of both sample and running buffer as input (also referred to as collection buffer or wash buffer). The array used a mirrored geometry to maximize concentration of collected product, with two arrays on either side of a common buffer stream. With only laminar flow in the array there is no turbulent mixing at the boundaries and diffusion is minimized by the ∼1 second residence time. The locations of the boundaries are dictated by the ratio of input fluids, and the offset pump heads on the Curate system help manage pulsatility, which leads to stable boundary locations. As cells are deflected by the array on either side of the channel they travel through the boundaries between the two streams and into clean running buffer in the central collection channel, shown in Figure 2C. Only cells and fluid in this central collection channel are collected as product, the remainder of fluid in the array goes to waste. A small portion of the running buffer is also rejected to waste to make sure that the collected product is free of diffusible species that may have crossed the boundary. This mode of operation is called Separation and Wash. An alternative mode of operation can increase the concentration of white blood cells in the product by recirculating the collected product back through the array as collection buffer. This is facilitated by a recirculation path on the cassette and easily controlled by actuating just 2 valves open/closed (Figure 1A, top). This mode of operation, titled Separation, Wash and Concentration, requires no additional processing time and the output concentration can be adjusted based on the downstream assay requirements.

Total system throughput and capacity were accomplished through massive parallelization. 34 lanes were fabricated in a single layer and were accessed by a common set of manifold channels, one for each of product, collection buffer, product and waste (Figure 2D). Two layers were bonded to a central manifold layer, and then two of these manifold units were connected with y-connectors. The result was 134 lanes across 4 microfluidic layers operating in parallel, with only 2 inputs and 2 outputs from the cassette. The throughput of the final geometry was 1000 mL/hr, with sample and collection buffer inputs comprising 400 mL/hr and 600 mL/hr respectively, and product and waste outputs generated at 300 mL/hr and 700 mL/hr respectively.

#### Performance of the DCS system

The cell sorting system processed undiluted apheresis samples at 400 mL/hr, resulting in fully automated processing times of under an hour with minimal hands-on time up front. Apheresis collections from both Terumo Spectra Optia™ and Fresenius Kabi Amicus™ systems were processed, and example collected products are shown in Figure 3A.

**Figure 3.**
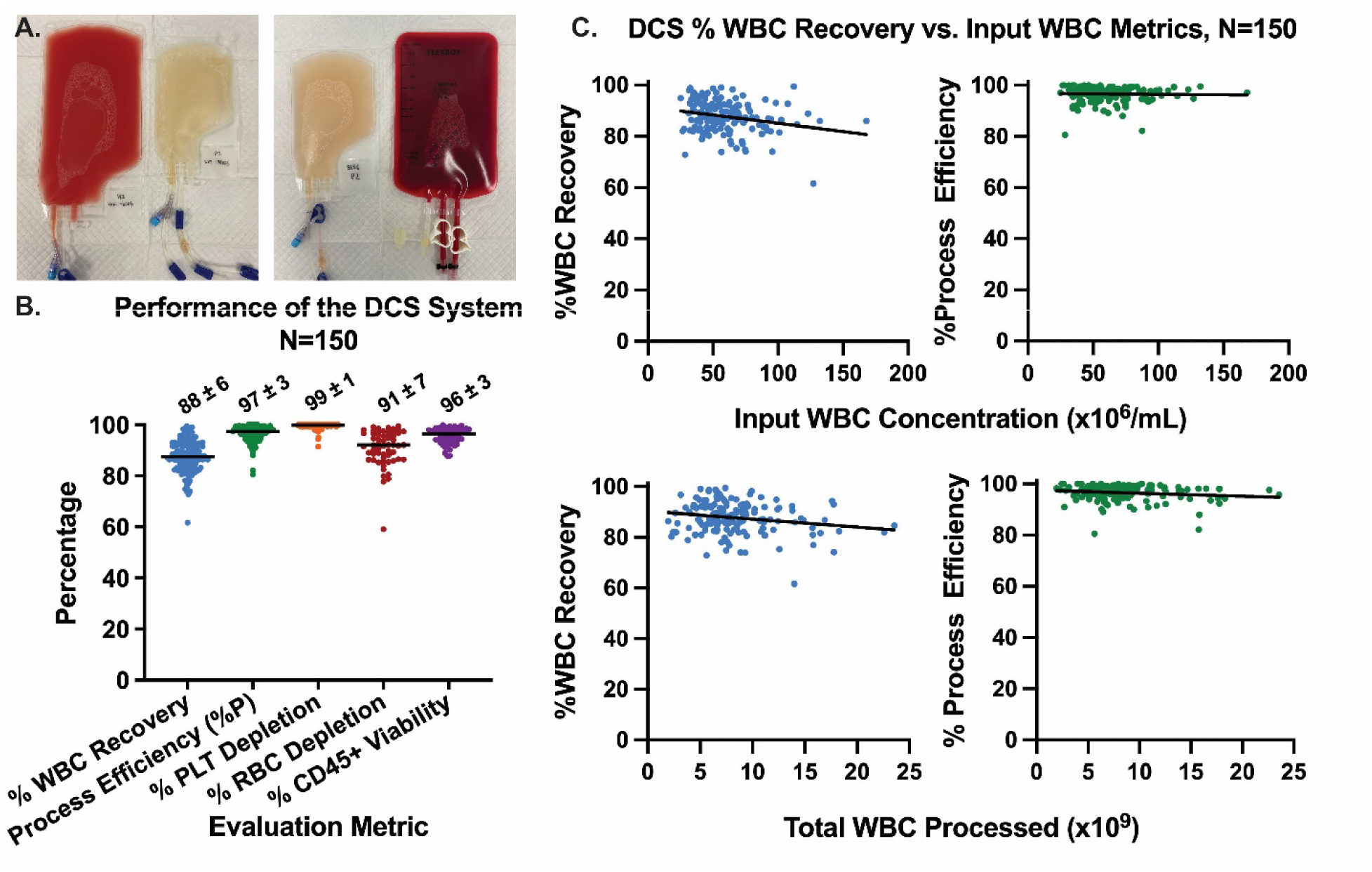
A) Example DCS products and wastes generated by processing apheresis samples. B) The performance of the DCS system across N=150 samples evaluated by %WBC depletion, % Process Efficiency, %PLT depletion, %RBC depletion, and %CD45+ viability. Values indicate mean +/-standard deviation and horizontal line indicates median value. C) %WBC recovery vs. input concentration (top) and vs. total WBCs processed (bottom). Calculated slopes are shown by the black lines. %WBC recovery plots both had a slight downwards trend, while the % Process efficiency showed no significant trend.

N=150 normal donor apheresis collections were processed, undiluted, on the system to demonstrate its performance across a wide range of donor material. The system’s in-line pressure detection upstream of the microfluidic array enabled an automated programmatic scaling of throughput should there be any issues detected, and without requiring any user intervention. Of the 150 samples loaded only 9 runs were auto scaled by the instrument, and of those runs, 7 ran to completion at the scaled flow rate of 240 mL/hr. The other 2 runs still processed >85% of the sample and recovered >85% of the total white blood cells (WBCs) loaded, while preserving the remainder of the sample to be processed on another cassette. There was no correlation between which samples triggered instrument autoscaling and input sample composition.

The performance metrics from all N=150 runs are shown in Figure 3B. The average % WBCs recovered was 88 +/- 6%. The array Process Efficiency (cells in the product vs. all collected cells in product and waste bags) was much higher, with 97 +/-3% of all cells recovered in the product. The unrecovered cells (not in product or waste) were likely trapped upstream of the microfluidic array by the in-line 20 µm pre-filter configured to remove cell clumps before they impacted array performance. The purity of the collected products was evaluated by measuring platelet (PLT) and red blood cell depletion (RBC). %PLT depletion was 99 +/1% and % RBC depletion was 91 +/- 7% respectively. Viability was determined by staining CD45+ cells with 7-AAD and was 96 +/- 3%, equivalent to the input population. No difference in performance between Separation and Wash vs. Separation and Wash with Concentration modes was observed (Supplemental Figure 1).

The key separation metrics of %WBC recovery and % process efficiency (product vs. waste) were evaluated for dependency on input concentration or on total cells processed. Surveying the N=150 samples, leukopack concentrations up to 168 M/mL (average of 56 +/- 24 ×10^6^ M/mL), total WBC cellularity up to 24 billion leukocytes (average 8.5 +/- 3.8 billion leukocytes) and volumes up to 245 mL (average of 150 +/- 50 mL) were processed without dilution or modification of protocol. While a slight decrease in %WBC recovery was observed with both increasing input concentrations and increasing input total WBCs processed, the % process efficiency showed no impact (t-test, deviation from 0 slope not significant). This indicates that with higher concentrations and/or total WBCs processed there were more cells trapped in the upstream filter, likely due to increased cell clumping. Material that passed the upstream filter was still separated with equivalent efficiency compared to lower concentrations and counts.

## Recovery and Depletion Performance of DCS vs. manual Ficoll® -Paque®

The DCS system processed full leukopacks within an hour. By comparison, processing the same volume by manual density-gradient centrifugation (Ficoll-Paque^®^) required up to 4h hours with significant hands-on time, increased contamination risks, and yielded operator dependent results. Performance of the system was tested against manual Ficoll® by processing the same set of leukopacks by each method, removing ∼10-20 mL for Ficoll® and processing the remainder via DCS. Recoveries of each method for N=86 split samples are shown in Figure 4.

**Figure 4.**
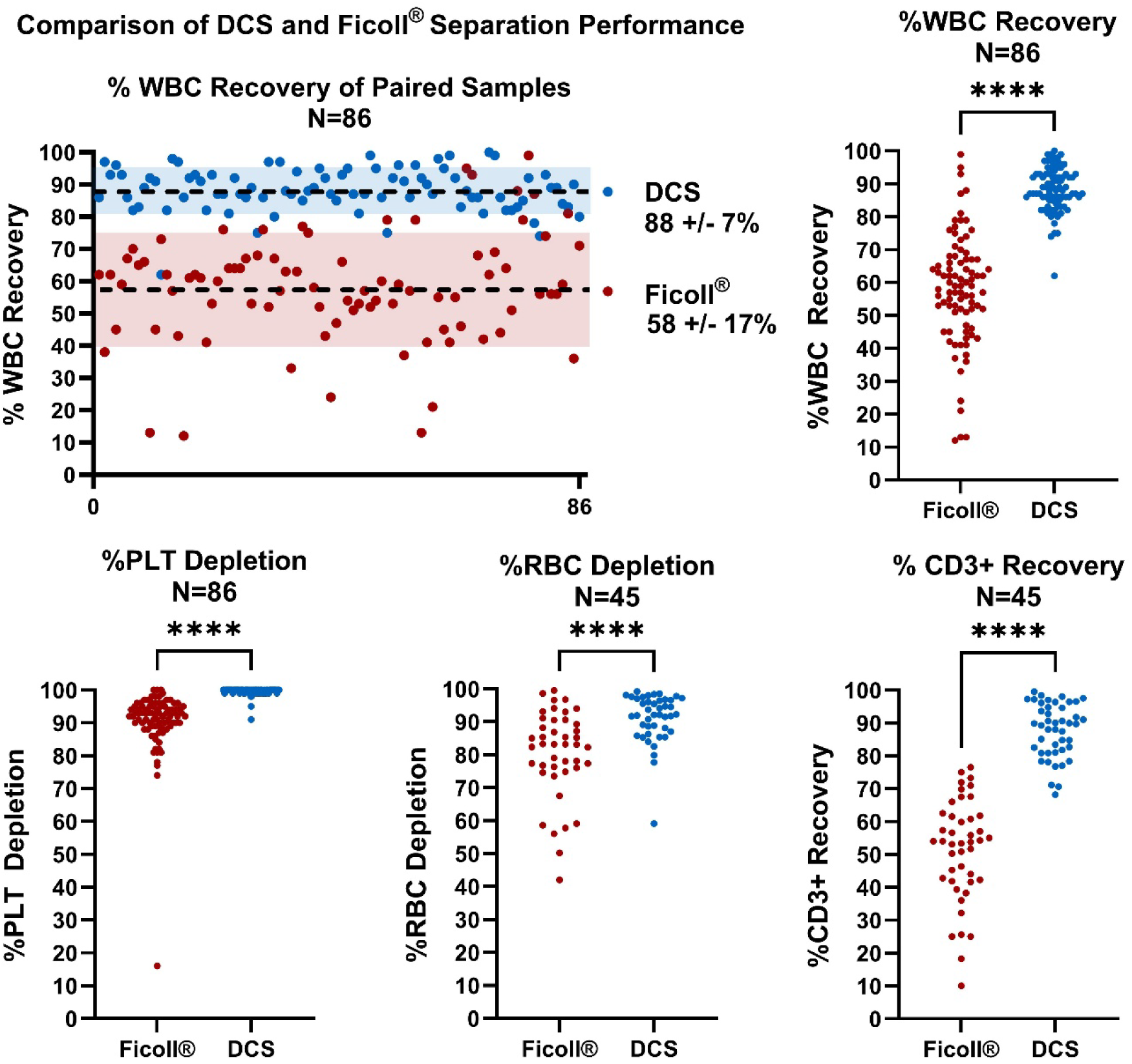
Paired processing metrics of DCS vs. Ficoll® preparation methods. % WBC recovery, % PLT depletion, % RBC depletion and % CD3+ recovery were all significantly higher in the DCS system. RBC depletion and CD3+ recovery were determined by staining a random subset (N=45) of the paired samples for FACS analysis. (****) = p<0.0001, all comparisons two-tailed t-tests. Horizontal lines indicate median values.

DCS recovered 88 +/-7% of WBCs loaded while Ficoll® only recovered 58 +/- 17%, indicating an average of ∼1.5x more cells recovered by the microfluidic DCS system. The DCS system also showed an improvement in washing efficiency compared to manual density gradient centrifugation (Figure 4, bottom left). The PLT depletion was ∼10 fold better and significantly more consistent in DCS-processed samples, with a % depletion of 99 +/-1% for the Curate system vs. 91 +/-10% for Ficoll-Paque®. A subset of samples was stained and analyzed by flow cytometry to better quantify residual RBC and CD3+ T-cell concentrations. Out of N=45 samples the RBC depletion was ∼2 -fold better in DCS than Ficoll®. The recovery of CD3+ T-cells showed similar trends to the %WBC recovery (Figure 4, bottom right), with ∼1.7x more CD3+ cells recovered by DCS than by Ficoll®.

### Relative Purity of DCS vs. manual Ficoll-Paque®

DCS delivered higher WBC recovery coupled with greater RBC and PLT depletion, resulting in a purer WBC population of cells. The ratios of RBC/WBC and PLT/WBC are shown in Figure 5A and B. Input ratios were calculated using Coulter Counter measurements prior to processing. Product ratios were determined by staining for the different cell types and analyzing by flow cytometry. The DCS system reduced the RBC/WBC ratio by 70-fold, a greater than 2-fold improvement over Ficoll®. There was a more significant difference when comparing the PLT depletion, with DCS-achieving an average ∼100-fold reduction in the PLT/WBC ratio, a ∼10 fold improvement over Ficoll®. The combined impact was an improvement in purity from on average ∼2% in the input material to only ∼14% via Ficoll®, but over 60% by DCS processing.

**Figure 5.**
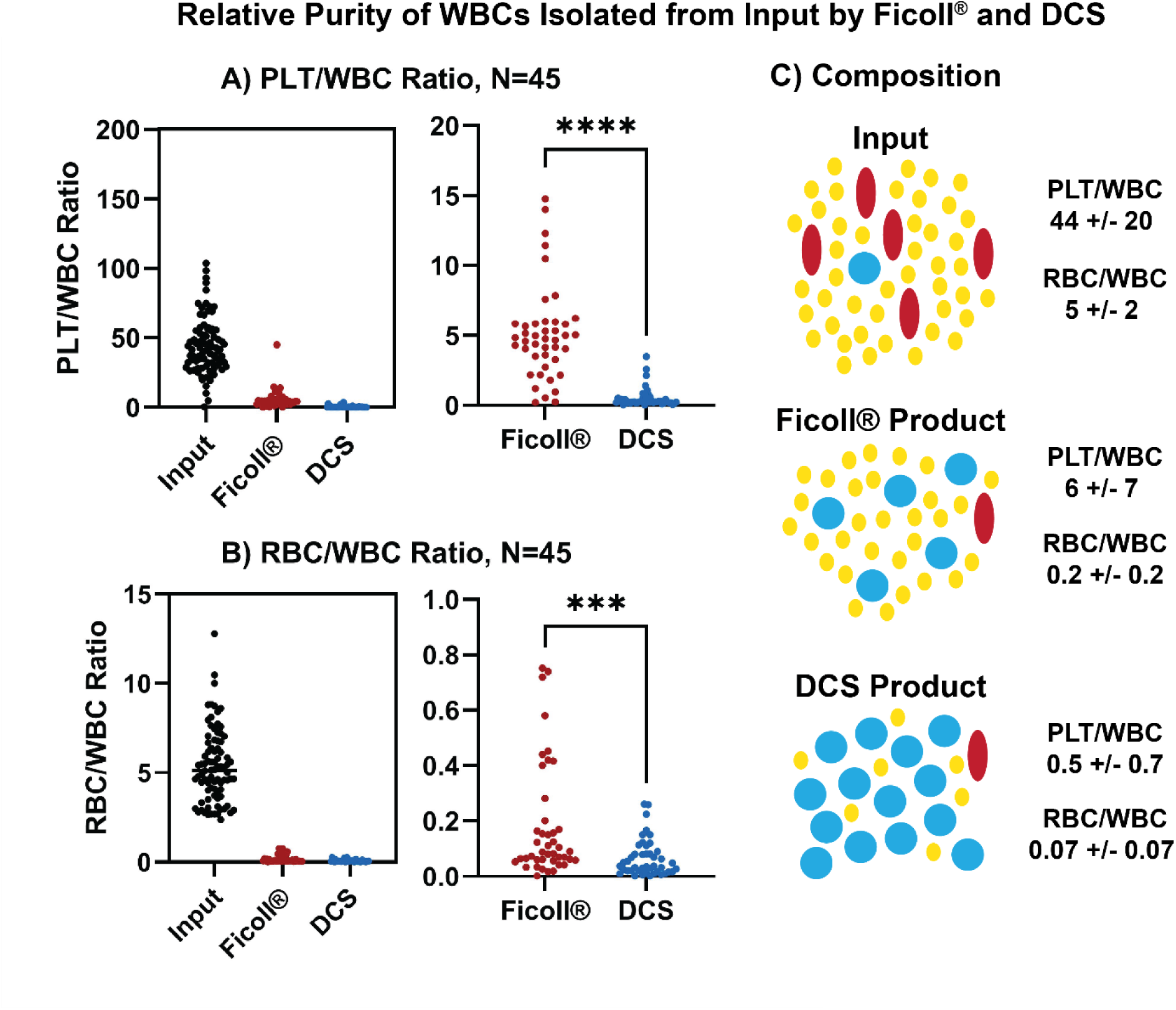
Purity of the cells collected via DCS or Ficoll® preparation. A) The DCS system yields ∼100x reduction in PLTs per WBC, vs. only ∼10x reduction for Ficoll® (p<0.0001, two-tailed t-test). B) The DCS system yields >10x reduction in RBCs per WBC vs. only ∼4x for Ficoll® (p=0.0004, two-tailed t-test). C) Composition of the average Input sample, Ficoll® product, and DCS product. Numerical values indicate average +/- standard deviation. The DCS system recovered more WBCs while depleting more RBCs and PLTs, resulting in fewer contaminating cells per WBC collected and an overall purer product.

### T-cell phenotype

Recent publications have shown that pre-activation negatively impacts the transformation to CAR-T cells, leading to loss of therapeutic potential.^14,15^ Recognizing potential process differences with respect to cell:cell interaction and tonic signaling, a subset of products (paired, N=21) were phenotyped by flow cytometry within two hours of processing. CD69+ was chosen as a marker to indicate activation levels in the cell population. Basal product activation state was measured prior to any cell engineering. As shown in Figure 6A, the Ficoll® prepared cells had on average ∼2x higher percentage of CD69+ T-cells (p=0.0018). The Ficoll® prepared cells also had more variability across the N=21 samples, with one product showing >25% of the CD3+ cells that were also CD69+.

**Figure 6.**
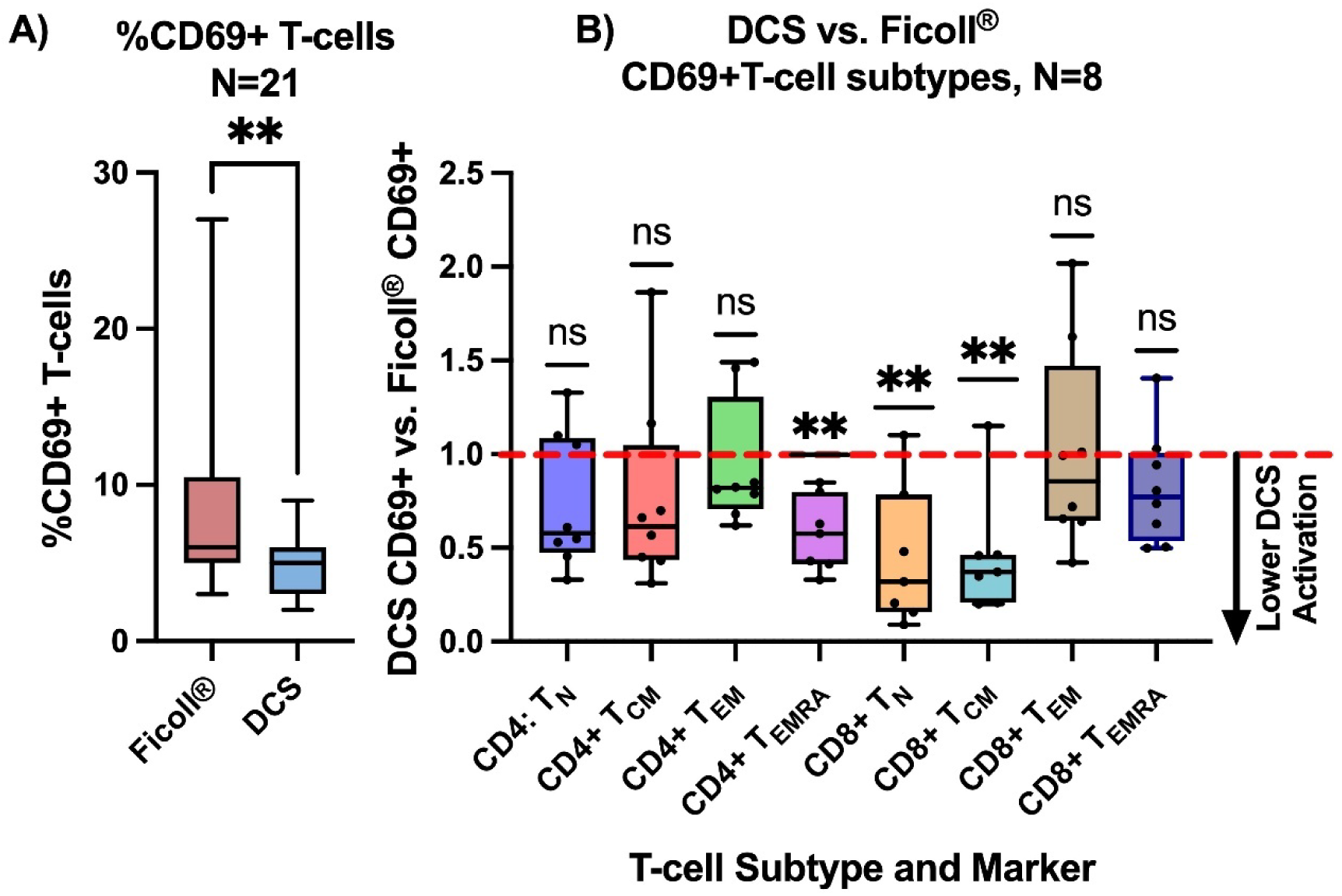
A) CD69+ activation of Ficoll® vs. DCS-purified cells. Ficoll® purified cells were ∼2x more activated, indicated by % of CD3+ cells that are also CD69+. Box and whiskers defined by interquartile ranges and min to max points. p=0.0018, two-tailed t-test. B) CD69+ activation within the T-cell subtypes, indicated as a ratio of DCS/Ficoll®. Ratios <1 indicate higher activation in the Ficoll®-purified cells. Box and whiskers defined by inter-quartile ranges and min to max points. Ratios for each T-cell type were compared against the theoretical value of 1 using a one-sample, two-tailed t-test (significance alpha=0.05). (**) = p<0.01, (ns) = not significant.

A smaller subset of samples (paired, N=8) were phenotyped to investigate CD69+ expression within the T-cell effector differentiation pathway subsets. The 4 major subtypes are naïve T-cells (T_N,_ CCR7+CD95-CD45RA+), central memory T-cells (T_CM,_ CCR7+CD95+CD45RA-CD62L+), effector memory T-cells (T_EM,_ CCR7-CD95+CD45RA-CD62L-), and terminally-differentiated effector memory (T_EMRA,_ CCR7-CD95+CD45RA+CD62L-).^43^ Each of these subtypes play a different role when engineered with a CAR-T, ranging from short-lived but potent effector function (T_EM_) to long-lived *in vivo* expansion and proliferation (T_CM_).^44^ The %CD69+ was determined within each subtype and the plot is shown as a ratio of DCS-prepared cells vs Ficoll®-prepared cells.

Equivalent %CD69+ activation is indicated by the red dotted line (value =1). In all subtypes analyzed a similar pattern was observed. The Ficoll® prepared CD4+ and CD8+ cells on average showed higher %CD69+ activation, resulting in mean ratio values <1. Using a one-sample, two-tailed T-test with a significance of 0.05 the individual ratios were compared to a theoretical value of 1. The CD4+ T_EMRA_, CD8+ T_N_ and CD8+ T_CM_ samples showed a ratio significantly different than 1, indicating that the DCS cells had lower activation within those subsets.

### Washing efficiency (Cytokines)

To understand a potential underlying cause of the phenotype differences at Day 0, cytokines from the DCS and Ficoll®-prepared product fractions were collected (N=2 samples analyzed in duplicate, collected within 2 hrs of purification). A full panel analysis indicated numerous cytokines present in the Ficoll® preparation at levels exceeding 50 pg/mL, generally considered to be substantial (Figure 7). In addition, several cytokines that are inhibitors of T-cell proliferation, including PDGF^45,46^ (AA, BB and AB/BB), and markers of cells bring driven into senescence (PAI-1^47^) were found at levels 5-10x higher in Ficoll® vs. DCS products. Some cytokines and chemokines in Ficoll® were found at levels equivalent to or higher than the starting material, indicating active generation of cytokines as opposed to incomplete washing. The input material also contained alarming levels of some cytokines and chemokines, emphasizing why an efficient, fast wash is a critical first step. The highly potent proinflammatory chemokine RANTES was found at a level almost 2000-fold higher than the accepted biologically active levels in the source apheresis material. Following processing, Ficoll® with two wash steps eliminated ∼75% as compared to the DCS processing which eliminated ∼95% of RANTES. While this is only a small N it highlights how the different cellular purification efficiencies combined with the degree of soluble factor elimination of potent biological response modifiers are potentially impacting activation, differentiation and response of T-cells -and also other cells present.

**Figure 7.**
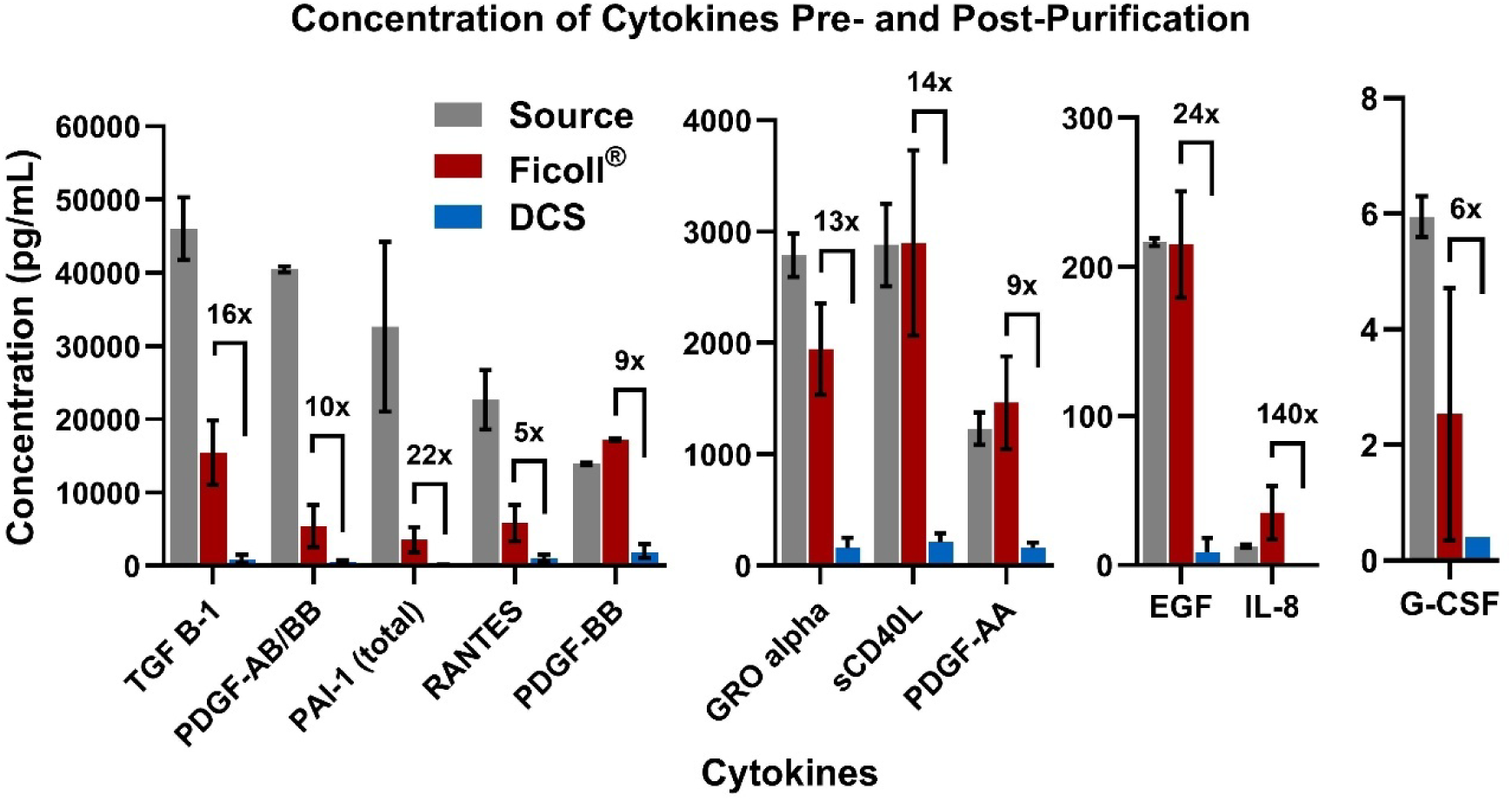
Concentration of cytokines present in the milieu both pre- and post-separation. N=2, samples analyzed in duplicate. Shown are the instances where the ratio of Ficoll® /DLD is >5x. For PAI-1, IL-8 and G-CSF some measurements from the DCS sample were below the assay’s limit of detection. Error bars indicate standard deviation.

### Expansion

A subset of the paired samples (N=20) was stimulated and expanded to observe expansion kinetics. The same number of T-cells were stimulated and plated for each head-to-head comparison of DCS and Ficoll®, and fold expansion was determined at days 3, 6, and 12. On average, by day 12 the DCS cells had expanded 14.7 fold vs. only 11.9 fold for Ficoll®, indicating almost 25% more cells in culture from DCS-purified starting material despite plating at the same density (Figure 8A). Using a two-tailed, paired T-test the fold expansion at Day 12 was found to be significantly different within the DCS and Ficoll populations (p<0.05). To demonstrate the impact of higher recovery from the DCS process, the starting value at day 0 was multiplied by the %CD3+ recovery ratio (DCS/Ficoll®) for each DCS sample (Figure 8B). Note that the average Ficoll %CD3+ recovery of 48 +/- 16% here is lower than the N=86 mean of 51.3 +/-16% but the difference between the populations is not significant. On day 0 the DCS cells began with 2.1-fold more CD3+ than Ficoll®, and by day 12 that difference grew to 2.6-fold more cells vs.

**Figure 8.**
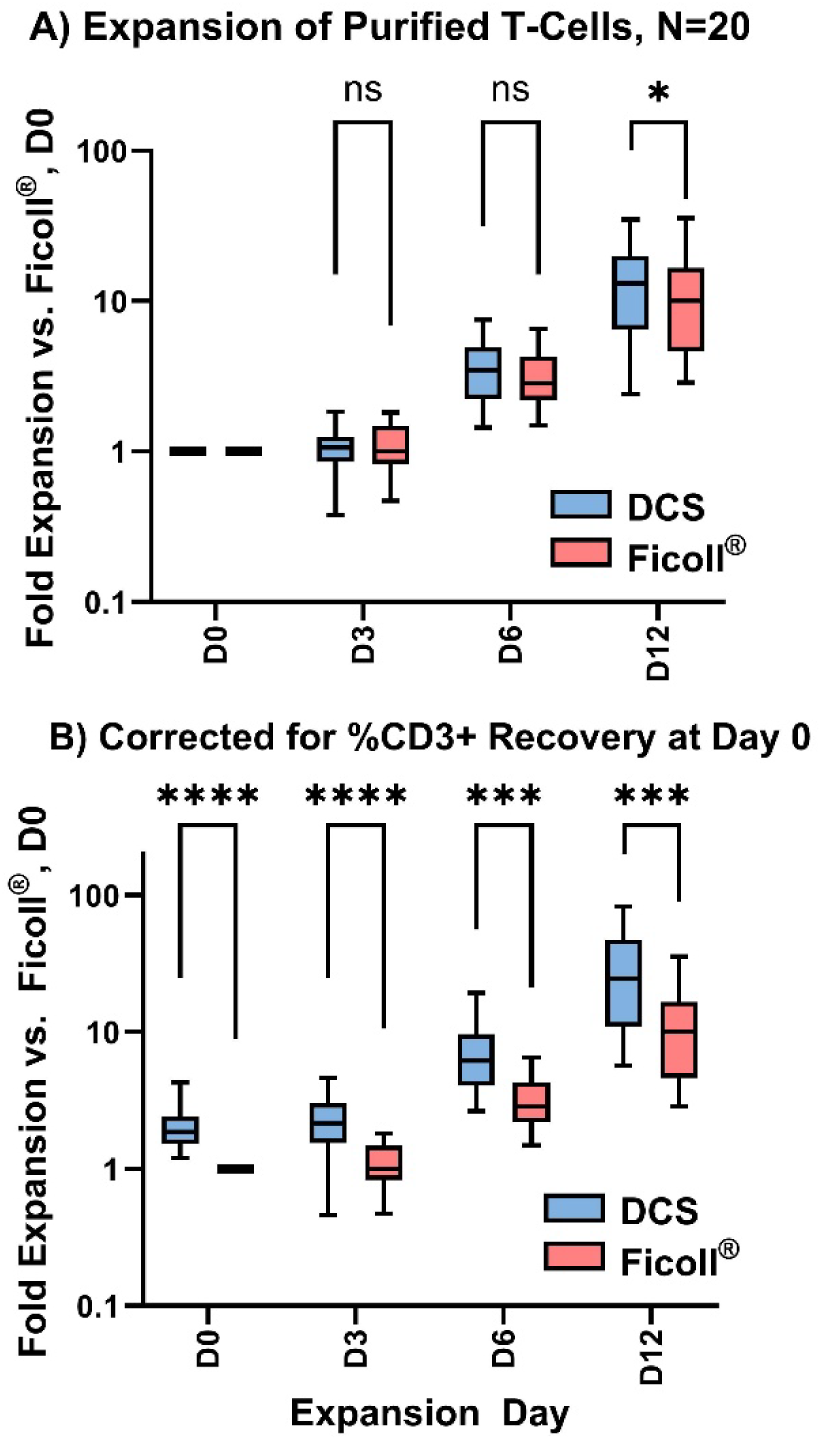
A) Expansion of CD3+ cells isolated from Ficoll® and DCS methods. At Day 12 cultures from DCS-purified cells had ∼25% more T-cells than Ficoll®. B) Fold expansion correcting for higher %CD3+ recovery of DCS at D0. Box and whiskers indicate inter-quartile range and min to max points. All comparisons are paired t-tests for that expansion day, two-tailed, significance = 0.05. (*) = p<0.05, (**) = p<0.01, (***) = p<0.001, (****) = p<0.0001, (ns) = not significant.

Ficoll®. The slightly higher expansion kinetics, compounded by the higher initial recovery and purity of the DCS method, resulted in the DCS method almost reaching the same number of cells at day 6-9 as Ficoll® reached on day 12.

## DISCUSSION

With several FDA-approved therapies already on the market, CAR-T cell therapy has emerged as a highly effective means of treating blood malignancies. Within the first commercial products, a 12–25-day (or longer) manufacturing process was routine, of which 7-15 days was used for the actual manufacturing process and the rest being QC and logistics. Even today, with shorter conceptual processes, multiple steps and processes are used on Day 0 to recover and purify T cells for subsequent selection and activation, which can lead to variable cell loss before the point of T-cell selection^48,49^.

Here we described DCS as a method to eliminate some of the inconsistencies and impurities while minimizing losses during the earliest stages of CAR-T cell manufacturing. Inherent to DCS, size-based processing is an exceptionally efficient washing process. The single-pass separation-wash process eliminates the cell losses and forced cell-cell interactions from repeated centrifugation and minimizes non-desirable activations coming from a variety of soluble factors.

In comparison with standard centrifugation methods, DCS is a minimally manipulative process that is faster and superior in terms of platelet and red blood cell reduction. Efficient platelet removal is key for CAR-T cell manufacturing where processes may induce changes in T cells and compromise their proper development.^29^ The %PLT removal and CD69+ activation data shown here supports that trend with ∼2x lower CD69+ activation in the DCS isolated T-cells. A smaller set of samples analyzed for T-cell subtypes also demonstrated a lower activation state in the DCS-prepared cells, with significantly less activation vs. Ficoll® demonstrated for CD4+ T_EMRA_, CD8+ T_N_ and CD8+ T_CM_ cells.

Furthermore, the efficient DCS wash process, in a small-scale study, resulted in lower levels of cytokines that negatively impact CAR-T therapies in the product. These cytokines have a range of known effects from inducing T-reg differentiation to suppressing T-cell expansion and proper generation of CAR-T. The exact cause of this reduction in cytokines for the DCS product could be the improved platelet depletion, the gentler process, or a combination of the two. This reduction in cytokines is particularly relevant for minimizing inflammatory responses and more severe cytokine release syndrome for shortened CAR-T processes with minimal (or no) expansion times.

## Conclusions

Our study is the first to demonstrate full-scale microfluidic processing of apheresis samples for CAR-T cell therapy. Besides the technological advances of a microfluidic system that can process ∼250 mL of apheresis sample in under an hour, our study shows that the inherent differences in the processing methods result in key differences in the purified products. In a field where centrifugation is the most widely used method to obtain cells, DCS is a novel, elegant and highly efficient approach to rapidly obtain demonstrably better starting material for downstream processing.

## Author Contributions

Alison M. Skelley, Yasna Behmardi, Khushroo Gandhi and Tony Ward designed, tested and optimized the Curate system and microfluidic arrays. Alison M. Skelley, Roberto Campos-Gonzales, and Tony Ward performed experiments, interpreted data, and prepared figures and the manuscript. Yasna Behmardi and Luke Petersen performed experiments and interpreted data. Mabel Shehada tested the Curate system and performed experiments. Laurissa Ouaguia performed experiments.

## Data Availability

All data produced in the present study are available upon reasonable request to the authors

## Acknowledgements

We are grateful to David Inglis for his assistance in contributing to the design of the microfluidic arrays. We are also grateful to Chris Boyce for his assistance in analyzing flow cytometry data and compiling summaries. Finally, we wish to acknowledge the thoughtful input from Dr. Curt Civin at University of Maryland Baltimore and Dr. James Sturm of Princeton University, both of whom were also funded by the STTR.

## Glossary

DCS: Deterministic Cell Separation
DLD: Deterministic Lateral Displacement
PBS: phosphate buffered saline
PLA: plasmalyte-A
LKP: leukopack
WBC: white blood cell
RBC: red blood cell
PLT: platelet
BSA: bovine serum albumin
HSA: human serum albumin
ACD-A: anticoagulant citrate dextrose solution, solution
A COP: Cyclic Olefin Polymer

