## Supplemental Information for "Improving Quality of CAR-T Cell Therapy Starting Material with Automated Microfluidic Cell Sorting"

**SUPPLMENTARY INFORMATION**

### Curate cassette and instrument

The cassette is a single-use, disposable, fully closed, sterile unit. Upon connection of the reagent, sample, and empty product and waste bags to the cassette, the user loads the cassette on the Curate system and hangs the bags on the corresponding load cells. Once the Curate door is closed, the system automatically processes the sample through a series of stages: DLD priming, run phase, and end of run product flush. The system monitors the run performance through a series of onboard sensors, including bubble sensors and pressure sensors upstream of the DLD, and load cells (hangers) from which fluids (buffer, sample, product, waste) are hung. Sensor feedback during the run phase is used to both monitor the run progress and to automatically take action to preserve the viability of the sample and fidelity of the DLD microarray. Bubbles detected upstream of the DLD are shunted to a DLD bypass, while an increase in pressure results in an automatic scaling of all the flow rates to keep processing conditions below 20 psi, preserving sample viability while also continuing to process. Bag masses are also used to queue certain script actions depending on the run mode selected.

### Mode of operation

Standard processing of a sample takes place in Separation and Wash mode. In this mode, sample and buffer are pumped into the DLD, and the target cells are deflected into the wash buffer and collected in the product channel. Cells smaller than the critical diameter are not deflected, and pass through the array into the waste channel. The final product characteristics (WBC concentration, volume) depend on the sample volume. Various running buffers can be used in this mode including albumin/ Plasma-Lyte®-A Injection, albumin/saline, common tissue culture medias such as Optmizer and TexMACs, and higher viscosity cryopreservation medias (up to 2 cP).

A modification of this mode is Separation and Wash with Concentration, a novel method of operating the DCS. This mode relies upon feedback from the product bag hanger to maintain a fixed product volume, as selected by the user at the start of the run. Once the target volume/mass is reached, the product is recirculated back into the cassette and DLD elements during the run, taking the place of the buffer input during cell sorting. The sample continues to process with target cells deflecting into the recirculated product stream. Up to ~5-fold concentration of the sample is possible, with collected volumes as low as 40 mL, while still achieving the recovery and depletion metrics achieved during standard modes of operation.

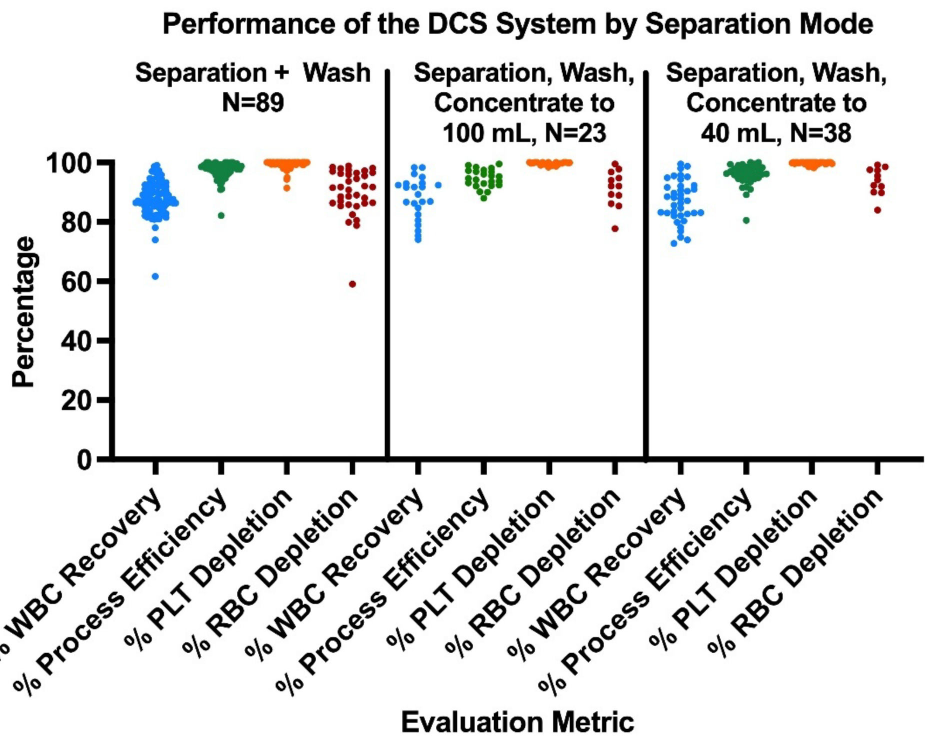


**Supplemental Figure 1:** Performance of the Curate system and DCS separation under different modes of operation. No difference in performance is observed between the 3 modes of operation.

### Cytokines

| Average of pg/mL*<br>Row Labels | Column Labels<br>Source | Ficoll | DCS | %REMOVED<br>FI/Source | DCS/Source | RELATIVE<br>Fi/DCS |
|---|---|---|---|---|---|---|
| Myeloperoxidase | 446,104.7 | 91,412.0 | 63,311.4 | 80% | 86% | 1.44 |
| Cathepsin D | 189,088.3 | 33,035.6 | 18,671.7 | 83% | 90% | 1.77 |
| PDGF-BB | 13,936.6 | 17,238.5 | 1,958.4 | -24% | 86% | 8.80 |
| TGF B-1 | 46,020.6 | 15,465.6 | 969.3 | 66% | 98% | 15.96 |
| NCAM | 206,721.4 | 10,658.4 | 4,755.5 | 95% | 98% | 2.24 |
| RANTES | 22,657.2 | 5,829.3 | 1,065.2 | 74% | 95% | 5.47 |
| PDGF-AB/BB | 40,493.6 | 5,451.5 | 550.8 | 87% | 99% | 9.90 |
| sICAM-1 | 39,261.9 | 4,381.7 | 2,992.3 | 89% | 92% | 1.46 |
| PAI-1 (total) | 32,623.8 | 3,565.9 | 164.3 | 89% | 99% | 21.71 |
| sCD40L | 2,881.4 | 2,897.9 | 209.9 | -1% | 93% | 13.80 |

| | | | | | | |
|---|---|---|---|---|---|---|
| GRO alpha | 2,788.0 | 1,943.1 | 155.3 | 30% | 94% | 12.51 |
| sVCAM-1 | 352,722.5 | 1,197.6 | 2,768.7 | 100% | 99% | 0.43 |
| PDGF-AA | 1,388.7 | 833.4 | 85.7 | 40% | 94% | 9.72 |
| BDNF | 8,248.9 | 470.6 | 141.2 | 94% | 98% | 3.33 |
| EGF | 216.8 | 215.0 | 8.8 | 1% | 96% | 24.45 |
| FGF-2 | 376.1 | 114.3 | 27.8 | 70% | 93% | 4.11 |
| MDC | 1,527.5 | 58.7 | 16.0 | 96% | 99% | 3.67 |
| VEGF-A | 223.0 | 52.8 | 22.4 | 76% | 90% | 2.36 |
| IP-10 | 182.4 | 38.1 | 25.4 | 79% | 86% | 1.50 |
| MIP-1B | 263.2 | 37.8 | | 86% | 100% | - |
| IL-8 | 12.7 | 35.1 | 0.3 | -175% | 98% | 140.30 |
| MCP-1 | 247.1 | 15.7 | | 94% | 100% | - |
| IFNa2 | 5.3 | 12.7 | 5.3 | -140% | 0% | 2.40 |
| Fractalkine | 5.6 | 9.9 | 4.0 | -77% | 29% | 2.49 |
| IL-1a | | 6.2 | 6.5 | - | - | 0.95 |
| TNFa | 26.9 | 4.5 | 3.3 | 83% | 88% | 1.37 |
| GM-CSF | 25.5 | 3.4 | 2.0 | 87% | 92% | 1.72 |
| G-CSF | 6.0 | 2.5 | 0.4 | 57% | 93% | 6.37 |
| IL-1RA | 702.3 | 1.8 | 0.5 | 100% | 100% | 3.88 |
| MIP-1a | 7.9 | 1.7 | 3.3 | 78% | 58% | 0.52 |
| IFNy | 1.9 | 1.0 | 0.2 | 49% | 88% | 4.34 |
| IL-12P70 | 2.4 | 0.7 | 0.6 | 70% | 74% | 1.15 |
| IL-17A | 2.5 | 0.7 | 0.6 | 73% | 77% | 1.16 |
| IL-3 | 0.6 | 0.6 | 0.6 | 9% | 6% | 0.97 |
| IL-1B | 1.4 | 0.4 | 1.1 | 69% | 22% | 0.39 |
| IL-7 | 3.6 | 0.4 | | 90% | 100% | - |
| IL-9 | 24.1 | 0.4 | 0.3 | 99% | 99% | 1.18 |
| IL-2 | 0.1 | 0.3 | 0.2 | -139% | -118% | 1.10 |

| | | | | | | |
|---|---|---|---|---|---|---|
| IL-4 | 34.4 | 0.2 | 0.4 | 99% | 99% | 0.55 |
| IL-5 | 0.1 | 0.0 | 0.0 | 80% | 94% | 3.33 |
| IL-12P40 | 34.0 | | | 100% | 100% | - |
| IL-15 | 2.7 | | | 100% | 100% | - |
| TNFB | | | | - | - | - |
| MCP-3 | | | | - | - | - |
| TGF-a | 2.0 | | 1.9 | 100% | 6% | - |
| Flt-3L | 33.3 | | | 100% | 100% | - |
| Eotaxin-1 | 220.1 | | | 100% | 100% | - |
| IL-18 | 861.3 | | 13.9 | 100% | 98% | - |
| IL-13 | | | | - | - | - |
| IL-6 | | | | - | - | - |
| IL-10 | 0.8 | | | 100% | 100% | - |

**Supplementary Table 1.** Cytokines present both pre- and post-separation by Ficoll and DCS. N=2 samples were analyzed in duplicate. Green boxes highlight all ratios >1 (Ficoll value greater than DCS). Highlighted green boxes with no value indicate that cytokines were detected in the Ficoll products, but in the DCS products they were not detected or below the range of linearity on the dilution curve. Yellow boxes: Ficoll product value was equivalent to or exceeded value from input material.
